# CD36 and SR-B1 Polymorphisms Exhibit Distinct Association Patterns in Active and Latent Tuberculosis

**DOI:** 10.1101/2025.11.21.25340627

**Authors:** Ezza Binte Tariq, Urooj Subhan, Farah Deeba, Zuha Tariq, Afrose Liaquat, Sidra Younis

**Affiliations:** Department of Biological Sciences, National University of Medical Sciences, Rawalpindi Pakistan; Department of Biochemistry, The Women University, Multan Pakistan; Department of Biosciences, COMSATS University, Islamabad Pakistan; Department of Biochemistry, Shifa College of Medicine, Islamabad Pakistan

**Keywords:** Scavenger receptors, rs1761667 (g.18436G>A), rs3211938 (g.73946T>G), rs4238001 (g.5275G>A), single nucleotide polymorphisms

## Abstract

**Introduction:** Host genetics plays a pivotal role in determining disease susceptibility among individuals infected with *Mycobacterium tuberculosis* (Mtb). Scavenger receptors (SRs) such as *CD36* and *SR-B1* mediate pathogen recognition and lipid uptake, both of which are central to mycobacterial entry and immune modulation.

**Gap Statement:** Polymorphisms rs1761667 and rs3211938 in *CD36* and rs4238001 in *SR-B1* have not been investigated in any population in relation to both latent tuberculosis infection (LTBI) and active TB.

**Aim:** To genotype *CD36* and *SR-B1* polymorphisms and evaluate their association with TB and LTBI. To predict functional/regulatory impact of these SNPs and compare their allele frequencies with global datasets.

**Methodology:** Polymorphisms were genotyped using ARMS-PCR within a case-control design. Genotype frequencies were compared using Fisher’s exact chi-square test. Functional and regulatory effects were predicted using PolyPhen-2 and RegulomeDB, while the 1000 Genomes database was used for population comparison.

**Results:** The homozygous AA genotype of *SR-B1* rs4238001 was strongly associated with active TB (p = 0.00), while the heterozygous GA genotype showed a protective association with LTBI (p = 0.00). For *CD36*, the homozygous GG genotype of rs3211938 was associated with protection against active TB (p = 0.02) but exhibited the opposite pattern in LTBI (p < 0.00). Moreover, the heterozygous GA genotype of rs1761667 was significantly linked to increased risk of LTBI (p = 0.00). In silico functional prediction classified rs4238001 as missense and rs3211938 as nonsense variant. Regulatory analysis indicated rs4238001 and rs1761667 affect transcription in TB-relevant tissues. Population analysis highlighted variation in allele frequencies across groups.

**Conclusion:** Polymorphisms in *SR-B1* and *CD36* show distinct associations with LTBI and TB, suggesting contrasting genetic influences on infection establishment and disease onset. These findings reveal a novel host genetic component of TB pathogenesis and warrant validation in larger, multiethnic cohorts.

## 1. Introduction

Tuberculosis (TB) is a global health concern caused by *Mycobacterium tuberculosis* (Mtb), a highly infectious bacterium. This bacterium can exist in a latent state or an active disease state resulting in latent TB infection or active disease, respectively (1). TB primarily spreads via airborne respiratory droplets expelled when infected individuals cough or sneeze. According to the WHO Global TB Report 2025, an estimated 1.23 million people died from TB in 2024. Globally, approximately one quarter of the population (22–25%) is estimated to harbor latent Mtb infection. After initial infection, the likelihood of progressing to active TB is greatest within the first two years, affecting roughly 5% of infected individuals (2). Pakistan is ranked fifth among the countries with the highest TB burden in the world. The mortality rate of TB in Pakistan is 34 deaths per 100,000 people per year (3).

The progression and outcome of TB infection are intricately influenced by the interplay of pathogen, host, and environmental factors. The development of active TB in only a fraction of infected individuals suggests that host genetics is a pivotal determinant of disease development (4). Previous studies have identified a significant association of genetic polymorphisms in human and experimental animal models on the outcomes of Mtb infection (5,6).

Scavenger receptors (SRs) are one of the pattern recognition receptors (PRRs) that recognize mycobacterial pathogen-associated molecular patterns (PAMPs). SRs belong to a class of cell surface transmembrane glycoproteins that play vital roles in the control of macrophages and innate immunity (7). SRs are categorized into 12 classes, with class B comprising *SR-B1*, *SR-B2*, and *CD36*. SRs are one of the various cell surface and intracellular receptors that mediate the uptake of bacteria (8). SRs interact with the lipoglycans and lipoprotein components of mycobacteria and facilitate the direct molecular interaction between the host and Mtb (9). Inflammatory cytokines are produced by the activation of SRs, which subsequently activate the macrophages (10). Macrophages kill Mtb through phagocytosis and induce an adaptive immune response.

Previous studies have supported the involvement of mutations in SR genes with multiple diseases including infectious diseases (11,12), cardiovascular diseases (13,14), metabolic disorders (15,16), and neurodegenerative disorder (17). Study reveals the association of polymorphism rs4238001 with reduced expression and impaired function of *SR-B1* gene (18). Similarly, the SNPs rs1761667 and rs3211938 were found to be associated with reduction in *CD36* expression (19). Such functional variants may therefore influence host–pathogen interactions by altering receptor-mediated lipid uptake and immune signaling.

To date, only two studies have explored associations between SR gene (MARCO) polymorphisms and TB, both conducted in Gambian and Chinese populations (20,21). In addition, *CD36* variants have been studied in pulmonary TB among the Chinese Han population (22), but these investigations focused on different *CD36* polymorphisms (rs1194182, rs10499859) and MARCO variants. Importantly, none of these studies have evaluated the functionally relevant *SR-B1* and *CD36* polymorphisms analyzed in our work, nor has any study been conducted in a South Asian population.

To address this gap, our study extends previous investigations of SR genetics by focusing on polymorphisms (rs1761667, rs3211938, and rs4238001) known to alter receptor expression and lipid uptake. By including both LTBI and active TB groups, we aimed to determine how these polymorphisms may influence infection establishment versus disease onset. Therefore, we investigated the association of *CD36* gene polymorphisms rs1761667 (g.18436G>A) and rs3211938 (g.73946T>G), and *SR-B1* gene polymorphism rs4238001 (g.5275G>A) with TB and LTBI in the Pakistani population.

## 2. Material and methods

All the experiments in this study were performed following the ethical guidelines and regulations of the NUMS Institutional Review Board, in compliance with the Declaration of Helsinki. Blood samples were collected after receiving informed consent from patients with recently confirmed TB and their contacts at the Nishtar Hospital Multan (NHM). Blood samples of TB contacts were also collected from laboratory workers dealing with TB patients from TEMAR Diagnostics Rawalpindi and Pride Laboratory Lahore. Samples from healthy controls were collected from Raza Laboratory, Nishtar Road Multan.

GeneXpert/AFB positive cases were included in the TB patients’ group of study. TB contacts consisted of the household contacts who did not have blood relation with TB patients, had been in contact with index case of pulmonary TB within the last 6 months and were asymptomatic (i.e., no cough, weight loss, fever, night sweats, lymphadenopathy, or any symptom / sign of active TB). For healthy controls the following inclusion criteria were considered: absence of TB symptoms and no recent contact with active TB patients. All study subjects were ≥ 16 years of age and those tested positive for HIV infection or pregnant/ breastfeeding women were excluded from the study (Supplementary Table S1 and Annex 3).

We used Epitools software to calculate the sample size for this case-control study and the following parameters were considered: TB prevalence in Pakistan, the expected odds ratio, 95% confidence level, and the desired statistical power. The calculated sample size was 350 study subjects including 150 healthy controls, 100 active TB patients, and 100 TB contacts. Detailed information regarding consent forms, questionnaires and ethical approval is given in supplementary material (Annex 1 and Annex 2).

The genomic DNA from blood samples (1-2ml) was extracted using the phenol-chloroform method. *SR-B1* and *CD36* polymorphisms were identified from the National Center for Biotechnology Information dbSNP database (https://www.ncbi.nlm.nih.gov/snp/). PRIMER 1 software was used to design primers for the SNPs rs4238001, rs1761667, and rs3211938. All SNPs were genotyped using amplification refractory mutation system PCR (ARMS-PCR). Details of the primers and PCR conditions are given in Supplementary Tables S2 and S3.

GraphPad Prism 10.0.2 software was used for statistical analysis. Age was compared between study groups using independent samples t-tests and one-way ANOVA. Normality was assessed using Shaprio-Wilk test, and homogeneity of variances was assessed using Levene’s test. Gender distribution was compared using the chi-square test. Fisher’s exact chi-square test was used to compare genotype and allele frequencies. Odds ratios (ORs) and 95% confidence intervals (CIs) were calculated for co-dominant, dominant, recessive, and additive genetic models. For all analyses, p-values < 0.05 were considered statistically significant.

We also conducted a thorough in silico analysis using different databases to further explore the functional implications of the studied SNPs. The potential effects of the SNPs on protein function were predicted using PolyPhen-2 via SNPnexus, and their regulatory impact was evaluated using RegulomeDB. Using the GTEx Portal, significant Expression Quantitative Trait Locus (eQTL) and Splicing Quantitative Trait Locus (sQTL) associations were investigated. To evaluate the distribution of the studied polymorphisms across different populations worldwide, allele frequencies were obtained from the 1000 Genomes database via Ensembl.

## 3. Results

### 3.1. Genotype frequency distribution

The mean ages for TB patients, TB contacts, and healthy controls were 36 ± 11, 42.5 ± 13, and 41 ± 13.2 years, respectively. Gender distribution across the groups (female/male) was 54/46 for TB patients, 47/53 for contacts, and 67/83 for healthy controls. Genotype patterns of the study groups were examined to investigate the association between the SNPs in the *SR-B1* and *CD36* genes and susceptibility or resistance to TB.

#### 3.1.1. Association of *CD36* (rs1761667) with active TB and LTBI

When the genotypes were compared between healthy controls and TB patients in the co-dominant model a statistically significant *p* value was obtained (*p*=0.03, OR=0.73, 95%CI=0.43-1.24). The frequency of GA was higher in healthy controls (66.89%) as compared to TB patients (55.20%). The genotype frequencies of GG and AA in healthy controls *vs.* TB patients had minor differences (Healthy controls: GG: 32.41% and AA: 0.01%; TB Patients; 39.58% and 0.05%,). The genotype comparison in the recessive model gave statistically significant *p* value (*p*=0.02, OR=0.12, 95%CI=0.01-0.94), while in the dominant model (*p*=0.25, OR=0.73, 95%CI=0.43-1.24) and additive model (*p*=0.76, OR=0.94, 95%CI=0.64-1.38) statistically non-significant *p* values were observed (Table 1a).

**Table 1.** Genotype comparison of *CD36* and SRB1 polymorphisms between healthy controls (HC) and (i) active TB patients (TP), and (ii) TB contacts (TC).

| Gene-SNP | Genetic models | Genotypes | HC n (%)<br>145 (100) | TP n (%)<br>96 (100) | OR<br>(95%CI)<br>HC vs TP | P<br>value | $\chi^2$<br>value | TC n (%)<br>97 (100) | OR<br>(95%CI)<br>HC vs TC | P<br>value | $\chi^2$<br>value |
| --- | --- | --- | --- | --- | --- | --- | --- | --- | --- | --- | --- |
| a. |  |  |  |  |  |  |  |  |  |  |  |
| CD-36<br>rs1761667 | Co-dominant | GG | 47 (32.41) | 38 (39.58) | 0.73 (0.43-1.24) | 0.03 | 6.84 | 12 (12.37) | 3.39 (1.73-6.58) | 0.00 | 12.67 |
|  |  | GA | 97 (66.89) | 53 (55.20) |  |  |  | 84 (86.57) |  |  |  |
|  |  | AA | 1 (0.01) | 5 (0.05) |  |  |  | 1 (0.01) |  |  |  |
|  | Dominant | GG | 47 (32.41) | 38 (39.58) | 0.73 (0.43-1.24) | 0.25 | 1.30 | 12 (12.37) | 3.39 (1.73-6.58) | 0.00 | 12.66 |
|  |  | GA+AA | 98 (67.59) | 58 (60.42) |  |  |  | 85 (87.63) |  |  |  |
|  | Recessive | AA | 1 (0.69) | 5 (5.21) | 0.12 (0.01-0.94) | 0.02 | 4.85 | 1 (1.03) | 0.66 (0.03-12.79) | 0.77 | 0.08 |
|  |  | GA+GG | 144 (99.31) | 91 (94.79) |  |  |  | 96 (98.97) |  |  |  |
|  | Additive | G | 191 (65.86) | 129 (67.19) | 0.94 (0.64-1.38) | 0.76 | 0.09 | 116 (57.43) | 1.43 (0.98-2.08) | 0.05 | 3.61 |
|  |  | A | 99 (34.14) | 63 (32.81) |  |  |  | 86 (42.57) |  |  |  |
| b. |  |  |  |  |  |  |  |  |  |  |  |
|  |  |  | HC n (%)<br>119 (100) | TP n (%)<br>99 (100) |  |  |  | TC n (%)<br>88 (100) |  |  |  |
| CD-36<br>rs3211938 | Co-dominant | TT | 10 (8.40) | 9 (9.09) | 0.91 (0.34-2.20) | 0.02 | 7.06 | 13 (14.77) | 0.52 (0.21-1.30) | <0.00 | 18.74 |
|  |  | GT | 67 (56.30) | 71 (71.71) |  |  |  | 23 (26.15) |  |  |  |
|  |  | GG | 42 (35.29) | 19 (19.19) |  |  |  | 52 (59.09) |  |  |  |
|  | Dominant | TT | 10 (8.40) | 9 (9.09) | 0.91 (0.34-2.20) | 0.02 | 7.06 | 13 (14.77) | 0.52 (0.21-1.30) | 0.14 | 2.07 |
|  |  | GT+GG | 109 (91.60) | 90 (90.91) |  |  |  | 75 (85.23) |  |  |  |
|  | Recessive | GG | 42 (35.29) | 19 (19.19) | 2.29 (1.22-4.19) | 0.00 | 6.95 | 52 (59.09) | 0.37 (0.21-0.66) | 0.00 | 11.56 |
|  |  | GT+TT | 77 (64.71) | 80 (80.81) |  |  |  | 36 (40.09) |  |  |  |
|  | Additive | T | 87 (36.55) | 89 (44.95) | 1.97 (1.28-2.98) | 0.07 | 3.16 | 49 (27.84) | 1.49 (0.97-2.29) | 0.06 | 3.48 |
|  |  | G | 151 (63.45) | 109 (55.05) |  |  |  | 127 (72.16) |  |  |  |
| c. |  |  |  |  |  |  |  |  |  |  |  |
|  |  |  | HC n (%)<br>110 (100) | TP n (%)<br>80 (100) |  |  |  | TC n (%)<br>89 (100) |  |  |  |
| SR-B1<br>rs4238001 | Co-dominant | GG | 40 (36.36) | 17 (21.25) | 2.18 (1.14-4.33) | 0.00 | 14.75 | 54 (60.67) | 0.37 (0.20-0.66) | 0.00 | 12.75 |
|  |  | GA | 66 (60.00) | 46 (57.51) |  |  |  | 34 (38.20) |  |  |  |
|  |  | AA | 4 (3.63) | 17 (21.25) |  |  |  | 1 (1.12) |  |  |  |
|  | Dominant | GG | 40 (36.36) | 17 (20.73) | 2.18 (1.14-4.33) | 0.01 | 5.50 | 54 (60.67) | 0.37 (0.20-0.66) | 0.00 | 11.67 |
|  |  | GA+AA | 70 (63.64) | 63 (79.27) |  |  |  | 35 (39.33) |  |  |  |
|  | Recessive | AA | 4 (3.63) | 17 (20.73) | 0.13 (0.04-0.40) | 0.00 | 14.62 | 1 (1.12) | 0.30 (0.02-1.87) | 0.26 | 1.26 |
|  |  | GA+GG | 106 (96.36) | 63 (79.27) |  |  |  | 88 (98.88) |  |  |  |
|  | Additive | G | 146 (66.36) | 80 (50.00) | 1.97 (1.28-2.98) | 0.00 | 10.29 | 142 (79.78) | 0.50 (0.31-0.78) | 0.00 | 8.84 |
|  |  | A | 74 (33.64) | 80 (50.00) |  |  |  | 36 (20.22) |  |  |  |

$\chi^2$ : chi square, OR: odds ratio CI: confidence interval. The significant *p* values are shown in italics.

#### 3.1.2. Association of *CD36* (rs1761667) with LTBI

When the genotypes were compared between healthy controls and TB contacts in the co-dominant model statistically significant *p* value was obtained (*p*=0.00, OR=3.39 95% CI=1.73-6.58). The frequency of genotype GG was higher in healthy controls (32.41%) as compared to TB contacts (12.37%) whereas the frequency of GA was higher in TB contacts (86.57%) as compared to healthy controls (66.89%). The frequency of AA was similar in both study groups. Genotype comparison in the dominant model (*p*=0.00, OR=3.39 95% CI=1.73-6.58) gave statistically significant *p* value, while in recessive (*p*=0.77, OR=0.66, 95%CI=0.03-12.79) and additive model (*p*=0.05, OR=1.43, 95%CI=0.98-2.08) statistically insignificant *p* values were obtained (Table 1a).

#### 3.1.3. Association of *CD36* (rs3211938) with active TB

When the genotypes were compared between healthy controls and TB patients in the co-dominant model, a statistically significant *p* value was obtained (*p*=0.02, OR=0.91 95%CI=0.34-2.20). The frequency of genotype GG was higher in healthy controls (35.29%) compared to TB patients (19.19%), whereas the frequency of genotype GT was higher in TB patients (71.71%) than in healthy controls (56.30%). The frequency of genotype TT was almost similar in both groups. The comparison of healthy controls and TB patients in the recessive model (*p*=0.00, OR=2.29, 95%CI=1.22-4.19) exhibited statistically significant *p* value, while in dominant (*p*=0.85, OR=0.91, 95%CI=0.34-2.20) and additive model (*p*=0.07, 0R=1.97, 95%CI=1.28-2.98) statistically non-significant *p* values were observed (Table 1b).

#### 3.1.4. Association of *CD36* (rs3211938) with LTBI

When the genotypes were compared between healthy controls and TB contacts in the co-dominant model a statistically significant *p* value was obtained (*p*<0.00, OR=0.91 95% CI= 0.34-2.20). The frequency of genotype GT was higher in healthy controls (56.30%) compared to TB contacts (26.15%), whereas the frequency of genotype GG was higher in TB contacts (59.09%) compared to healthy controls (35.29%). The frequency of TT was similar in healthy controls and TB contacts. The comparison of genotypes in the dominant (*p*=0.00, OR=0.91 95% CI= 0.34-2.20) and additive model (*p*=0.00, OR=1.49, 95%CI=0.97-2.29) showed statistically significant *p* values, whereas the recessive model yielded a statistically insignificant *p* value (*p*=0.26, OR=0.37, 95%CI=0.21-0.66) (Table 1b).

#### 3.1.5. Association of *SR-B1* (rs4238001) with active TB and LTBI

When the genotypes were compared between healthy controls and TB patients in the co-dominant model significant *p* value was obtained (*p*=0.00, OR=2.18 95% CI=1.14-4.33). The frequency of GG was higher in healthy controls (36.36%) compared to TB patients (21.25%), whereas the frequency of genotype AA was higher in TB patients (21.25%) than in healthy controls (3.63%). The frequency of genotype GA was not different between healthy controls (60.00%) and TB patients (57.51%). When genotypes were compared in dominant (*p*=0.00, OR=2.18 95% CI=1.14-4.33), recessive (*p*=0.00, OR=0.13, 95% CI=0.04-0.40), and additive models (*p*=0.00, OR=1.97, 95% CI=1.28-2.98) statistically significant *p* values were obtained (Table 1c).

#### 3.1.6. Association of *SR-B1* (rs4238001) with LTBI

When the genotypes were compared between healthy controls and TB contacts in the co-dominant model a statistically significant *p* value was obtained (*p*=0.00, OR=0.37 95% CI=0.20-0.66). The frequency of genotype GG was higher in TB contacts (60.67%) compared to healthy controls (36.36%), whereas the frequency of genotype GA was higher in healthy controls (60.00%) than in TB contacts (38.20%). The difference in frequency of genotype AA between healthy controls (3.63%) and TB contacts (1.12%) was not significant. Genotype comparison in dominant (*p*=0.00, OR=0.37 95% CI=0.20-0.66), and additive model (*p*=0.00, OR=0.50, 95% CI=0.31-0.78) gave statistically significant *p* values, while in recessive model (*p*=0.26, OR=0.30, 95% CI=0.02-1.87) statistically insignificant *p* value was obtained (Table 1c).

### 3.2. In Silico Functional Annotation

An in silico functional analysis was conducted on the SNPs rs1761667, rs3211938 and rs4238001 to complement the genotypic association findings.

#### 3.2.1. Functional Impact and Regulatory Potential Prediction

Variant effect prediction using PolyPhen-2 classified the missense SNP rs4238001 of *SR-B1* gene as probably damaging with a high confidence score of 0.999, suggesting a potential alteration in *SR-B1* protein function. The SNP rs3211938 was predicted by SNPnexus to cause loss of protein while rs1761667, a non-coding mutation, is not expected to affect the protein structure. RegulomeDB analysis indicated a strong regulatory impact of rs1761667 and rs4238001, supported by transcription factor binding and expression data, with both SNPs assigned a RegulomeDB rank of 1f. However, rs3211938 showed minimal regulatory evidence with an assigned rank of 6. Table 2 summarizes the predicted functional and regulatory characteristics of the three studied SNPs. The transcription factor binding evidence is based on RegulomeDB annotations. All three SNPs are in actively transcribed chromatin regions, supported by DNase hypersensitivity and histone modification data.

**Table 2.** The table summarizes data from RegulomeDB and PolyPhen-2 (via SNPnexus) for both functional impact and regulatory potential prediction.

| Feature | rs1761667 | rs4238001 | rs3211938 |
| --- | --- | --- | --- |
| **Mutation Type** | Promoter SNP (non-coding, regulatory) | Missense (Gly2Ser) | Stop-gain (nonsense) |
| **Functional Impact** | Not protein-altering | Alters protein | Truncates protein |
| **Regulatory Effect** | Alters transcription factor binding & *CD36* expression | Affects SR-BI function and gene regulation | No strong regulatory evidence |
| **Transcription factor Binding** | POLR2A, CTCF, FOS | POLR2A, CTCF, JUN, RAD21 | POLR2A, CTCF, JUN, BCL3* |
| **Chromatin State** | Actively transcribed | Actively transcribed | Actively transcribed |
| **TB Relevance** | May impair phagocytosis & lipid uptake | May disrupt cholesterol transport & immune signaling | May cause immune dysregulation via protein loss |

\*indicates weak experimental evidence. POLR2A: RNA Polymerase II Subunit A, CTCF: CCCTC-Binding Factor, FOS: FBJ Murine Osteosarcoma Viral Oncogene Homolog, JUN: Jun Proto-Oncogene, RAD21: Cohesin Complex Component, BCL3: B-Cell Lymphoma 3 Protein

#### 3.2.2. Gene Expression and Splicing Effects

eQTL analysis using GTEx revealed that rs3211938 was significantly associated with decreased *CD36* expression in lungs and whole blood tissues. sQTL analysis indicated that rs1761667 was associated with significant alternative splicing changes in lungs, whole blood tissues, and spleen. These findings are shown in Figure 1.1 & 1.2. No significant eQTL or sQTL associations were found for rs4238001.

**Figure 1.1.**
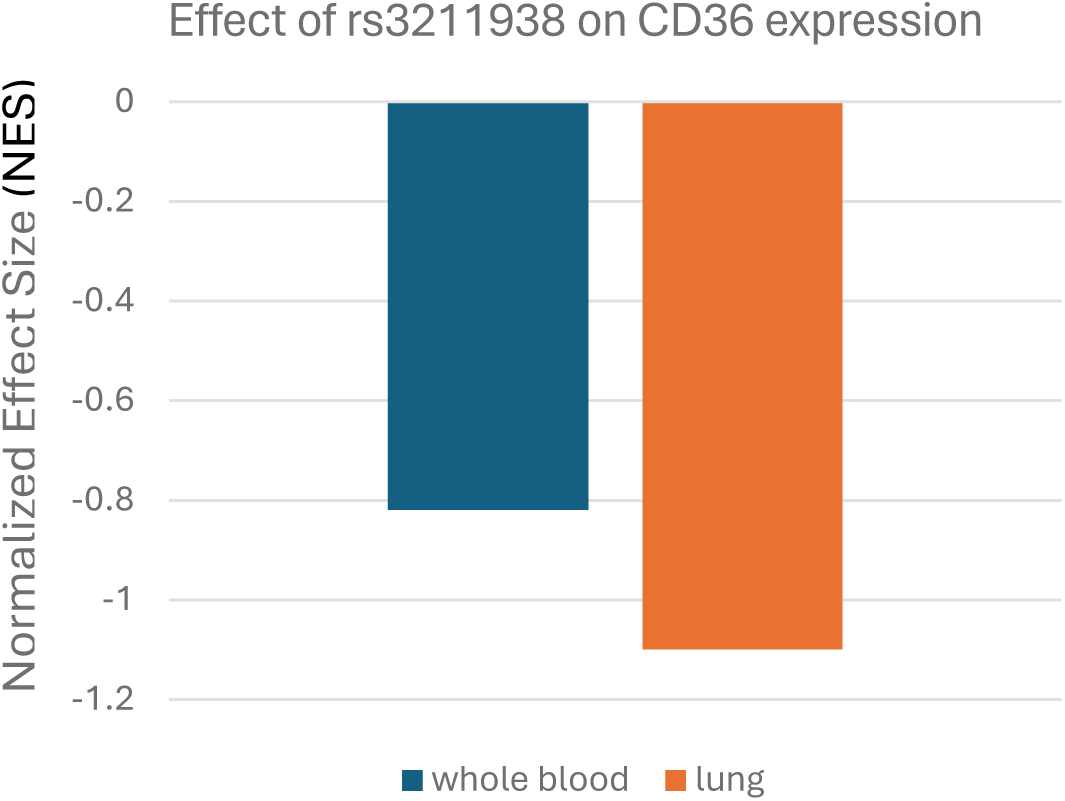
Bar graph showing the normalized effect size (NES) of the rs3211938 SNP on CD36 gene expression in whole blood and lung tissues based on GTEx expression quantitative trait loci (eQTL) analysis. The negative NES value indicates that the minor G allele of this SNP is associated with significantly decreased gene expression in both tissues (p = 2.07e−19 in whole blood, p = 3.94e−9 in lung).

**Figure 1.2.**
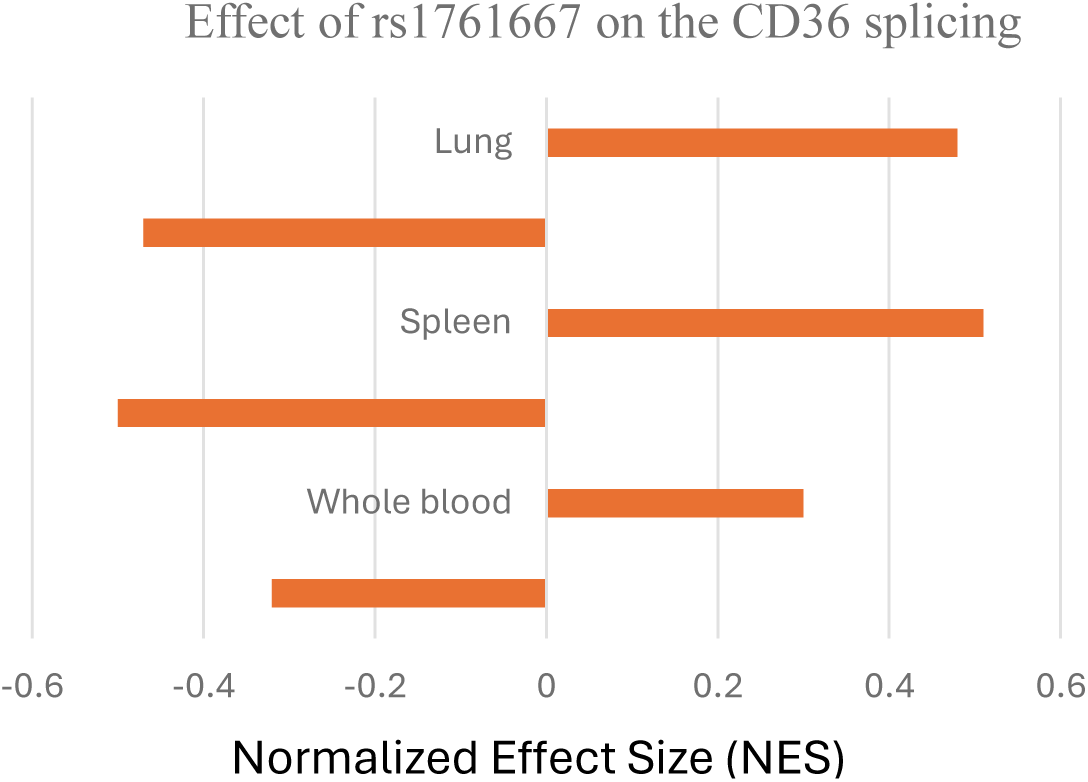
Bar graph representing the normalized effect size (NES) of rs1761667 on CD36 splicing events (sQTL) in whole blood, lung, and spleen tissues. The presence of both positive and negative NES values in the same tissue indicates allele-specific effects on different splicing events within the gene. Data retrieved from GTEx.

#### 3.2.3 Population Genetics Analysis

To determine the prevalence of the studied SNPs in various populations, Minor allele frequencies (MAF) were obtained from 1000 Genomes database. The promoter SNP rs1761667 was the most widely distributed, with the highest frequency in Europeans and Americans (53%), followed by Africans (35%), East Asians (31%), and South Asians (29%). The SNP rs3211938 was observed at a notable frequency of 12% in Africans but was absent in other populations. The missense SNP rs4238001 showed moderate frequency across several populations, with the highest MAF in Europeans (12%), followed by Americans (10%), Africans (7%), and South Asians (5%), while it was absent in East Asians. These findings indicate population-specific differences in SNP prevalence, supporting the need for population-focused genetic studies. The distribution patterns are illustrated in Figure 2.

**Figure 2.**
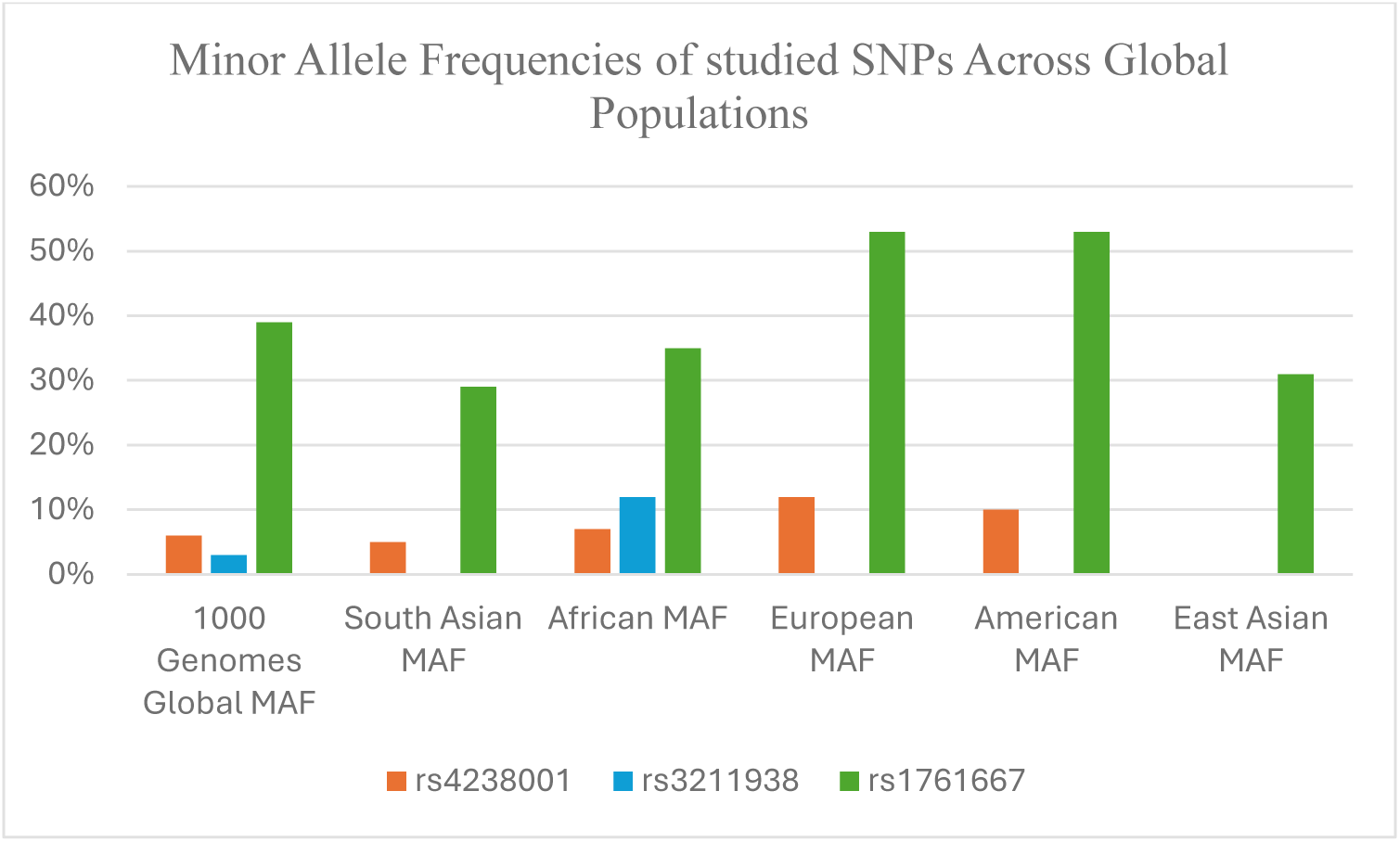
Bar chart showing the minor allele frequencies (MAF) of rs4238001, rs3211938, and rs1761667 in six global populations based on 1000 Genomes Project Phase 3 data.

## 4. Discussion

Host genetic factors play a significant role in determining susceptibility or resistance to Mtb infection. In our study, we focused on two scavenger receptors, *SR-B1* and *CD36*, which are part of the host’s innate immune system. We investigated the association between SNPs in these genes, namely rs4238001 in *SR-B1* and rs1761667 and rs3211938 in *CD36*, and TB in the Pakistani population. Our findings revealed a significant association between rs3211938 and rs1761667 in *CD36* and rs4238001 in *SR-B1* with active TB and LTBI. These results contribute to our understanding of the genetic factors involved in susceptibility of TB and may have implications for future research.

We found that the mutant genotype GG in rs3211938 was associated with resistance against active TB (p=0.02, OR=0.91 95%CI=0.34-2.20), as the frequency of mutant genotype was higher in healthy controls (35.29%) compared to TB patients (19.19%). The SNP rs3211938 (T>G), located on Exon 10, introduces an amino acid change from Tyrosine to termination codon at position 325 in *CD36* protein which affects the expression and function of the protein. Consistent with this, in silico prediction by SNPnexus indicated loss of function, while GTEx analysis revealed significantly reduced *CD36* expression in lungs and blood. The ‘G’ allele of rs3211938 is associated with reduced *CD36* expression and provides protection against atherogenic profile (19). Mechanistic studies further indicate that *CD36* is markedly upregulated in macrophages infected with Mtb and facilitates the accumulation of intracellular lipid droplets that provide a carbon source for bacterial persistence. Reduced expression of *CD36* therefore limits lipid uptake and foam-cell formation, restricting bacterial growth within macrophages (23). We hypothesize that a decrease in expression level may result in reduced mycobacterial growth as *CD36* is involved in the uptake of surfactant lipids by macrophages which promotes the growth of Mtb within macrophages (24). This mechanism potentially contributes to protection against TB. Our results are consistent with a previous study conducted on the association of *CD36* and *MARCO* with PTB in the Chinese Han population, indicating the significant association of SNPs in *the CD36* gene with resistance to PTB (22). While Lao et al. report the association in a Chinese Han cohort, our results describe association of functional polymorphisms in *CD36* in Pakistani population, conferring that population-specific genetic variation may influence the *CD36*-mediated susceptibility to TB. Our research findings are further supported by an *in vivo* study conducted on mice, where mice lacking the *CD36* gene showed resistance against mycobacteria. The absence of *CD36* led to a reduced ability of mycobacteria to survive within the cells. The study also indicated that *CD36* plays a role in cellular processes associated with the formation of granulomas, which aid in the initial growth and spread of bacteria (25). Therefore, we can conclude that the protective role of the GG mutant genotype in rs3211938, which is linked to a decrease in *CD36* expression, subsequently contributes to the inhibition of Mtb growth and spread.

Interestingly, the genotype GG of rs3211938 showed an opposing trend in comparison of healthy controls *vs.* TB contacts (p<0.00, OR=0.91 95% CI= 0.34-2.20) group. The frequency of genotype GG was higher in TB contacts (59.09%) compared to healthy controls (35.29%). Il’in and Shkurupy reported increased *CD36* expression in multi-nuclear phagocytes during Mtb persistence in BCG-infected mice (26). This could help explain why TB contacts show higher GG frequency and thus susceptibility to latent infection. These results may indicate a dual role of *CD36*, where decreased expression protects against progression to active TB by limiting lipid availability for bacterial replication, yet may favor the establishment of latent infection by impairing bacterial clearance during the initial phase. This interpretation is supported by recent evidence showing that lipid droplets act as multifunctional organelles that Mtb exploits during latency to evade immune clearance (27). Hence, a single gene might play different roles in determining susceptibility or resistance to LTBI and active TB. Our findings also align with the study investigating the association of the *SP110* gene with active TB and LTBI in Taiwan, which revealed the differential role of the gene for active TB and LTBI (28).

In rs1761667, we found a higher frequency of heterozygous genotype GA in TB contacts (86.57%) than healthy controls (66.89%) suggesting a significant association of rs1761667 GA genotype with risk to LTBI (p=0.00, OR=3.39, 95% CI=1.73-6.58). The SNP rs1761667 is located at Exon 1A, a G>A nucleotide change has been associated with lower *CD36* expression (19). Supporting this, GTEx splicing QTL analysis showed significant alternative splicing in lungs, whole blood, and spleen that are key immune tissues in TB. These results align with the study conducted in the Chinese Han population to investigate the association of *CD36* with carotid atherosclerosis. The results suggested the risk of disease in female patients carrying the GA genotype at rs1761667 (29). Another study suggested a strong association of the GA genotype with an increased risk of coronary heart disease in the Chongqing Han population of China (30). The *CD36* gene is responsible for mediating the effects of Mannose-capped lipoarabinomannan (ManLAM), leading to the release of TNF-α in peritoneal murine macrophages. Ligands of SRs have similar effects on TNF-α, and NO production as observed with ManLAM (31). *CD36* also facilitates lysosomal enzyme transportation and internalizes mycobacteria (32). Given its involvement in both lipid metabolism and pathogen recognition, reduced *CD36* expression due to rs1761667 may not only impair cytokine release but also alter intracellular lipid handling, potentially creating a microenvironment more favorable for latent infection. Moreover, *CD36* is a downstream target of the PPARδ signaling pathway, which integrates lipid metabolism and immune regulation. Activation of PPARδ promotes lipid droplet formation, a mechanism recently linked to Mtb persistence (33). Polymorphisms like rs1761667 that lower *CD36* expression could thus alter PPARδ-driven lipid–immune crosstalk, affecting the host’s ability to control latent infection. We found a weak *p* value for the comparison of rs1761667 genotypes between healthy controls and active TB patients (p=0.03, OR=0.73, 95% CI=0.43-1.24). The difference in genotype frequency distribution between studied groups may not remain significant by increasing sample size. We conclude that rs1761667 at *the CD36* gene may be important in determining the risk of LTBI but not active TB.

Interestingly, we found that *SR-B1* SNP at rs4238001 was significantly associated with active TB (p=0.00, OR=2.18, 95% CI=1.14-4.33). The frequency of mutant genotype AA was higher in active TB patients (21.25%) compared to healthy controls (3.63%). The SNP rs4238001 (G>A), located at Exon 1, introduces an amino acid change from Glycine to Serine and was predicted by PolyPhen-2 to be probably damaging (score:0.999), suggesting substantial disruption of *SR-B1* protein. This amino acid change is associated with a decrease in *SR-B1* expression (18). *SR-B1* engulfs Mtb in mesenchymal stem cells (MSCs), which exhibit innate control of mycobacterial replication through autophagy. MSCs are found in both human and mouse Mtb granulomas and play an important role in TB pathogenesis (34). This SNP at rs4238001 may affect the phagocytosis of Mtb in MSCs and result in an impaired control of mycobacterium. *SR-B1* influences cellular lipid traffic and membrane cholesterol homeostasis, which in turn modulate host–pathogen interactions (35). Collectively, these findings suggest that functional disruption by rs4238001 may not only impair lipid handling but also alter early host–pathogen communication, potentially contributing to increased TB susceptibility observed in our cohort. This SNP has not been studied in TB, LTBI, or any other lung disease, however, two independent research groups reported significant association of rs4238001 with coronary heart disease and low progesterone level (36,37).

In the comparison of *SR-B1* SNP at rs4238001 between healthy controls *vs.* TB contacts, we found that heterozygous GA genotype was significantly associated with protection against LTBI (p=0 00, OR=0.37, 95% CI=0.20-0.66). *SR-B1* on microfold cells (M cells) interact with Mtb EsxA enabling it to cross airway mucosa and initiate infection. Disruption in *SR-B1* genes decreases Mtb binding and translocation across M cells (38). Overexpression of *SR-B1* in macrophages increases Mtb and BCG binding (39). We hypothesize that changes in *SR-B1* gene expression due to mutation may affect Mtb binding, resulting in protection against LTBI. Our results are consistent with the study conducted by Acton et al., in which the SNP rs4238001 was associated with protection towards atherogenic lipid profile in white men (40).

Previous genetic investigations of SRs in TB have mainly focused on other *CD36* loci and MARCO variants. However, none of the functional variants analyzed in this study (rs3211938, rs1761667, rs4238001) were examined. Notably, rs3211938 is a well-characterized nonsense variant that markedly alters *CD36* expression in eQTL and exome datasets, yet prior disease-association studies have focused primarily on cardiometabolic phenotypes. Similarly, rs1761667 has been linked to lipid metabolism and cardiovascular traits, but its potential role in TB susceptibility remains unexplored.

To support these genotype associations, we performed pathway enrichment analysis *via* Reactome which demonstrates the functional involvement of SRs in TB pathogenesis. *CD36* and *SR-B1* are involved in immune signaling pathways including host response to Mtb, escape mechanisms of Mtb from phagocytes, and modulation of host immune defenses during latent infection (Figure 3). This strengthens the hypothesis that genetic polymorphisms affecting these receptors could modulate susceptibility to active and latent TB infection.

**Figure 3.**
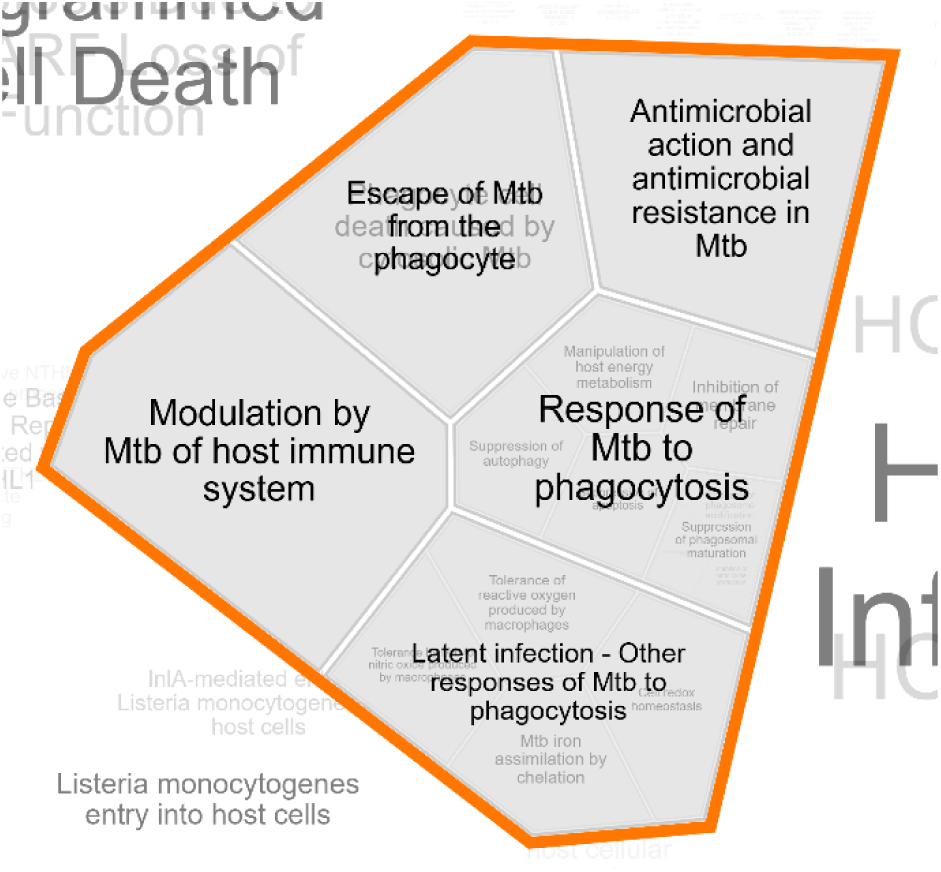
Reactome pathway enrichment map indicating the involvement of CD36 and SR-B1 in TB-related immune processes.

Additionally, our population genetic analysis from the 1000 Genomes database revealed significant ethnic variation in allele frequencies. For example, rs3211938 was only found in African populations, and rs1761667 had the highest frequency in Europeans and Americans. This supports the idea that population-specific studies are essential for understanding TB susceptibility. The distinct allele distribution observed here underscores the importance of examining functionally relevant receptor polymorphisms in high-burden populations like Pakistan, where host genetic diversity and environmental exposure may jointly shape TB risk.

Therefore, while our findings align with the broader concept that SRs influence TB susceptibility, this study is, to our knowledge, the first to assess these specific functional variants (rs3211938 and rs1761667 in *CD36*; rs4238001 in *SR-B1*) in the context of TB and LTBI, and the first to report such associations in a South Asian cohort. The integration of genetic association, in-silico functional predictions and pathway enrichment linking these receptors to immune signaling further supports their biological relevance.

## 5. Conclusion

In summary, this study is, to our knowledge, among the first to associate functional polymorphisms (rs3211938, rs1761667, rs4238001) in SR genes with TB outcomes, and the only report from a South Asian population. By integrating genetic association and in-silico analyses, our findings highlight the novelty and biological relevance of receptor-mediated lipid handling in TB pathogenesis.

While our sample size was statistically justified, it was relatively modest, which may limit generalizability to other ethnic groups. Moreover, LTBI classification was based on contact history rather than Interferon-Gamma Release Assay (IGRA) or QuantiFERON testing, and ARMS-PCR, though cost-effective, may not detect low-frequency mutations.

Future studies should therefore validate these findings in larger, ethnically diverse cohorts using sequencing-based technologies, employing functional assays to quantify receptor protein expression, and performing lipidomic profiling of infected macrophages. Such integrative approaches could further clarify how SR variation shapes host-pathogen interactions across different stages of TB infection.

## Data Availability

All data produced in the present work are contained in the manuscript

## Acknowledgements

We acknowledge the study subjects for providing their samples, the authors of the manuscript for their efforts, and the National University of Medical Sciences, Pakistan.

## Data Statement

All data generated during this study are included in this manuscript and its supplementary material. Additional data may be made available from the corresponding author upon request.

## Ethical Approval

This study was approved by the NUMS Institutional Review Board and conducted in accordance with the Declaration of Helsinki. Written informed consent was obtained from all participants.

*No new sequencing data were generated or analyzed in this study*.

