## Supplementary file for "CD36 and SR-B1 Polymorphisms Exhibit Distinct Association Patterns in Active and Latent Tuberculosis"

#### Supplementary Tables

**Table S1.** Inclusion and exclusion criteria applied while selecting study subjects.

| Study Group | Inclusion criteria | Exclusion Criteria |
|---|---|---|
| **Active TB patients** | Newly diagnosed with TB | HIV infected*<br>Pregnant/breastfeeding women* |
| **TB Contacts** | Household contacts<br>Asymptomatic | Clinical suspicion of active TB<br>Blood relation with active TB patient |
| **Healthy Controls** | Doesn't show symptoms of TB | Diagnosed with TB<br>Have been in contact with TB patient/s |

*These criteria were applied to exclude participants from all study groups. The age limit for all participants was ≥16 years.

**Table S2.** Sequence and properties of primers.

| Gene | Mutation | Primer Sequence | Length (bp) | Tm (°C) |
|---|---|---|---|---|
| ***SR-B1*** | rs5888 | **Outer forward:** ACACCCAAGTTCAAATCTGGCGTTGC | 26 | 66.0 |
| | | **Outer reverse**: ATGGAGTCGGGGTGAAGTAAGGAACT | 26 | 66.6 |
| | | **Inner forward:** CTCTCCCATCCTCACTTCCTCAACTCC | 27 | 68.8 |
| | | **Inner reverse:** CTTCTGCCAGAACCGGGGCA | 20 | 69.1 |
| | rs4238001 | **Outer forward:** CTCCCGCCCCAAAACGGAA | 19 | 67.7 |
| | | **Outer reverse**: CAGCAGCCCCTCCCGAAGC | 19 | 70.6 |
| | | **Inner forward:** CCAGGCGCGCAGACAGGA | 18 | 68.9 |
| | | **Inner reverse:** GCGCTTTGGCGGAGCATCC | 19 | 67.0 |
| ***CD36*** | rs1761667 | **Outer forward:** AAGGTCTGGTATCCACCTGTTTTCCT | 26 | 64.7 |
| | | **Outer reverse**: AAGAGTTTTCATGAAGCTTCCCGC | 24 | 61.9 |
| | | **Inner forward:** TTTTATTCATCTTTGCATGCCATCG | 25 | 57.3 |
| | | **Inner reverse:** TCATACTCCAGGCTTTGAGCATTGT | 25 | 61.9 |
| | rs3211938 | **Outer forward:** GAATAGTTCATGCTTGGCTATTGAGTTT | 28 | 58.2 |
| | | **Outer reverse**: CACCATTCTTTCTTCTGCCCTAATTACT | 28 | 60.4 |
| | | **Inner forward:** AAAATTATCTCAAAAAATTGTACATCACAG | 30 | 53.5 |
| | | **Inner reverse:** TGCATTTGCTGATGTCTAGCACATCA | 26 | 61.4 |

**Table S3.** PCR conditions for all single nucleotide polymorphisms.

| Mutation | PCR conditions | | | | |
|---|---|---|---|---|---|
| | **Initial denaturation** | **Denaturation** | **Annealing (35 cycles)** | **Extension** | **Final extension** |
| **rs5888** | 95 °C for 5 min | 95 °C for 1 min | 68 °C for 1 min | 72 °C for 1 min | 72 °C for 10 min |
| **rs4238001** | 95 °C for 5 min (for both first 10 cycles & 25 cycles) | 95 °C for 1 min (for both first 10 cycles & 25 cycles) | 65 °C (-1°C for every first 10 cycles) & 55 °C (for 25 cycles) 1 min | 72 °C for 1 min (for both first 10 cycles & 25 cycles) | 72 °C for 10 min |
| **rs1761667** | 95 °C for 5 min | 95 °C for 1 min | 61 °C for 1 min | 72 °C for 1 min | 72 °C for 10 min |
| **rs3211938** | 95 °C for 5 min | 95 °C for 1 min | 51 °C for 1 min | 72 °C for 1 min | 72 °C for 10 min |

**Annex 1. Ethical Approval**

NUMS
NATIONAL UNIVERSITY OF MEDICAL SCIENCES

National University of Medical Sciences
The Mall Rawalpindi
No: 06 / IRB&EC / NUMS / 24

18 Oct 2022

**CERTIFICATE OF APPROVAL - NUMS-IRB & ETHICAL COMMITTEE**

A research project titled "Scavenger Receptors Genes Association with Active and Latent Tuberculosis in Pakistani Population" in respect of PI-Dr Sidra Younis, Assistant Professor, Department of Biological Sciences, NUMS is hereby considered to be exempted for review by the IRB & Ethical Committee, NUMS.

Assistant Director ORIC
Secretary IRB & Ethical Committee
**(Aqeel Yunus)**

**COUNTERSIGNED**

Maj Gen
Pro-Vice Chancellor (Acad)
Chairman, NUMS-IRB & Ethical Committee
**{Saleem Ahmed Khan, HI(M) (Retd)}**

To: NDBS (For PI – Dr Sidra Younis)

**Project Title:** Association of Scavenger Receptor Genes Polymorphisms with TB in the Pakistani Population

**Organization:** National University of Medical Sciences, Rawalpindi, Pakistan

**Researcher:** Ezza Binte Tariq

1. **Reason for Conducting the Research:**
   TB is an infectious disease caused by bacteria (germs). TB infection is often inactive. Current tests for inactive TB are not suitable. We are trying to develop better diagnostic tests for inactive TB infection.
2. **Research Procedure:**
   You have been selected because you have been diagnosed with TB. If you provide written consent to participate in this study, you will be asked to provide details of your TB treatment. You will be asked to give a 5 mL blood sample for research tests, including a new blood test for TB infection. The entire process, including the consent form, filling out the questionnaire, and taking the blood sample, will take a maximum of 5 minutes. You will not need to return to the hospital. The blood test is for research purposes only.
3. **Benefits or Risks of Participation:**
   Rarely, giving blood may cause fainting. Very rarely, the site where blood is taken may develop an infection.
4. **Potential Benefits:**
   You will not receive direct benefit from participating in scientific research. This study may help in developing better diagnostic tests for inactive TB infection, which may benefit others in the future.
5. **Available Treatment for Adverse Events:**
   We do not expect any serious risks from participating in scientific research. To avoid any harm from dizziness, you will be asked to lie on a couch while giving blood. The sample will be taken by a trained member of the staff, who will clean the skin with an antiseptic.
6. **Confidentiality:**
   Your identity will be kept confidential in this scientific research. The study results, including laboratory or other data, may be published for scientific purposes, but your name will not be shown.
7. **Non-Participation in Research:**
   Taking part in scientific research is completely voluntary. It is up to you to decide whether to participate. Even after signing the consent form, you are free to withdraw from the study at any time without giving any reason. If you decide not to participate or withdraw from the study, your routine medical care will not be affected.
8. **Sources of Information:**
   For any questions related to the study and/or your rights, you may contact the researcher of the study.
   **Name:** Ezza Binte Tariq (Contact Info: 0304-0646405;)

**Patients Signature:**
**Patient's name:**
**Witness' signature:**
**Researcher's signature:**

I have read and understood the research work. I willingly agree to participate in this research study.

**Annex 3.** **Questionnaires:**

#### TB genetics Pakistan CRF: Active TB patients

##### 1. PERSONAL DETAILS

First name

Last name

Address

Mobile number

**Alternative contact:**

FAMILY MEMBER / FRIEND THAT MAY BE CONTACTED

Contact person/relationship

Contact person mobile number

##### 2. STUDY VISIT DETAILS AND ELIGIBILITY

Date of visit ______________________________________________

**Eligibility:**

| **Inclusion criteria** | **Yes** | **No** |
|---|---|---|
| Aged 16 or above | | |
| Newly diagnosed active TB about to start treatment | | |
| Gives written informed consent to participate in the study | | |
| **Exclusion criteria** | | |
| Known HIV infection | | |
| Pregnant or breastfeeding | | |

**Note**:

* If the answer to any of the inclusion criteria is **NO**, the subject is not eligible for the study *

**If the answer to any of the exclusion criteria is **YES**, the subject is not eligible for the study**

#### 3. DEMOGRAPHIC AND CLINICAL DETAILS

| | |
|---|---|
| Age | |
| Gender | |
| Ethnic origin | |
| Site of disease | |
| Basis of diagnosis (Smear, GeneXpert, other) | |
| Duration of symptoms | |
| Organism isolated | |
| Drug-susceptibility | |
| Previous anti-TB treatment | |
| Details if so | |
| Past medical history | |
| Current medications | |
| Smoking history | ○ Current ○ex-smoker ○never |
| BCG status (BCG scar present or absent?) | |

**4. RESULTS**

| **Laboratory test done** | **Result** |
|---|---|
| Haemoglobin level if available [state N/A if not available] | |
| Chest x-ray if available [state N/A if not available]<br>Date: | |

**Actual volumes (mL) of blood collected:**

| Tube | Target volume | Actual volume |
|---|---|---|
| EDTA | 5 ml | |

Date and time of blood collection: ________________________________________

Date and time of receipt of blood in laboratory: ______________________________

Received by (name of laboratory personnel): ________________________________

**Details of person completing CRF:**

Name ___________________ Signature ___________________________________

### TB genetics Pakistan CRF: Healthy controls

#### 1. PERSONAL DETAILS

First name

Last name

Address

Mobile number

**Alternative contact:**

FAMILY MEMBER / FRIEND THAT MAY BE CONTACTED

Contact person/relationship

Contact person mobile number

#### 2. STUDY VISIT DETAILS AND ELIGIBILITY

Date of visit ______________________________________________

**Eligibility:**

| **Inclusion criteria** | **Yes** | **No** |
|---|---|---|
| Aged 16 or above | | |
| Doesn't have symptoms of TB | | |
| Gives written informed consent to participate in the study | | |
| **Exclusion criteria** | | |
| Is diagnosed with TB/ has been in contact with TB patient | | |
| Known HIV infection | | |
| Pregnant or breastfeeding | | |

**Note**:

* If the answer to any of the inclusion criteria is **NO**, the subject is not eligible for the study *

**If the answer to any of the exclusion criteria is **YES**, the subject is not eligible for the study**

#### 3. DEMOGRAPHIC AND CLINICAL DETAILS

| | |
|---|---|
| Age | |
| Gender | |
| Ethnic origin | |
| Previous anti-TB treatment | |
| Details if so | |
| Past medical history | |
| Current medications | |
| Smoking history | ○ Current ○ex-smoker ○ever |
| BCG status (BCG scar present or absent?) | |

#### 4. RESULTS

| **Laboratory test done** | **Result** |
|---|---|
| Haemoglobin level if available [state N/A if not available] | |
| Chest x-ray if available [state N/A if not available]<br>Date: | |

**Actual volumes (mL) of blood collected:**

| Tube | Target volume | Actual volume |
|---|---|---|
| EDTA | 5 ml | |

Date and time of blood collection: ________________________________________

Date and time of receipt of blood in laboratory: ____________________________

Received by (name of laboratory personnel): ________________________________

**Details of person completing CRF:**

Name ___________________ Signature ___________________________________

**TB genetics Pakistan CRF: TB Contacts**

#### 1. PERSONAL DETAILS

First name

Last name

Address

Mobile number

Alternative contact:

FAMILY MEMBER / FRIEND THAT MAY BE CONTACTED

Contact person/relationship /

Contact person mobile number

#### 2. STUDY VISIT DETAILS AND ELIGIBILITY

Date of visit ___________________

**Eligibility:**

| **Inclusion criteria** | **Yes** | **No** |
|---|---|---|
| Aged 16 or above | | |
| Household contact with index case of pulmonary TB within the last 6 months | | |
| Asymptomatic (*i.e.,* no cough, weight loss, fever, night sweats, lymphadenopathy, or any other symptom / sign of active TB) | | |
| Gives written informed consent to participate in the study | | |
| **Exclusion criteria** | | |
| Clinical suspicion of active TB | | |
| Blood relation with active TB patient | | |
| Pregnant or breastfeeding | | |
| Known HIV infection | | |

**Note**:

* If the answer to any of the inclusion criteria is **NO**, the subject is not eligible for the study *

**If the answer to any of the exclusion criteria is **YES**, the subject is not eligible for the study*

**3. DEMOGRAPHIC AND CLINICAL DETAILS**

| | |
|---|---|
| Age | |
| Gender | |
| Ethnic origin | |
| Occupation | |
| Place of contact with index pulmonary TB case | ○ Household<br>○ Other |
| Index case's relationship with the participant | |
| Participant's proximity to index case | ○ Sleep in the same bed<br>○ Sleep in different bed but same room<br>○ Sleeps in a different room, same house<br>○ Sleeps in a different house |
| Activities shared with the index case | ○ Every day, >=50% of the day<br>○ Every day, <50% of the day<br>○ Not every day |
| How long was the index case coughing before their anti-TB treatment was started? | |
| Past medical history | |
| Current medications | |
| Smoking history | ○ Current ○ ex-smoker ○never |
| BCG status (BCG scar present or absent?) | |

**4. RESULTS:**

| **Test done** | **Result** |
|---|---|
| Haemoglobin level if available [state N/A if not available] | |

**Actual volumes (mL) of blood collected:**

| **Tube** | **Target volume** | **Actual volume** |
|---|---|---|
| EDTA | 5 ml | |

Date and time of blood collection: ________________________________________

Date and time of receipt of blood in laboratory: ______________________________

Received by (name of laboratory personnel): ________________________________

**Details of person completing CRF:**

Name ___________________ Signature ________________________________
